# Estimating the prevalence of late-onset Fabry disease in the US in 2024

**DOI:** 10.1101/2024.12.13.24319001

**Authors:** James Cook, Timothy Coker, Joshua Card-Gowers, Laura Webber

**Affiliations:** HealthLumen Limited, London, UK

## Abstract

Fabry disease is a rare lysosomal storage condition in which sphingolipid levels build up to harmful levels in various bodily organs, eventually leading to life-threatening complications such as stroke and kidney failure. Fabry disease is caused by rare pathogenic alleles in the *GLA* gene on chromosome X and may present as an early or late-onset disease depending on the identity of the causal allele and the severity of its effect on the gene product. Epidemiological studies have widely varied in their estimation of Fabry disease prevalence: estimates based on reported clinical cases range from 1 in 40,000 to 1 in 170,000 individuals, whilst recent estimates based on newborn screening are much higher, ranging from 1 in 1,250 to 1 in 21,973 individuals.

The primary aim of this study was to estimate the prevalence of Fabry disease in the US in 2024 by analysing selected *GLA* variants mostly associated with late-onset Fabry disease, projecting their allele frequencies to the US population and applying penetrance data from the literature to calculate how many causal allele carriers would be expected to be symptomatic for the disease at some point within their lifetime.

8 causal genetic variants were selected for analysis in this study based on their inclusion in a previous Fabry disease study using data from the UK Biobank. Allele frequencies for all 8 variants in global ancestry groups were extracted from gnomAD v4.1. The size and demographic makeup of the US population in 2024 was obtained from the US Census Bureau and mapped to gnomAD v4.1 ancestry groups, using previously reported estimates of the ancestral composition of Census groups encompassing multiple ancestry groups. Carrier counts by sex and ethnic group were calculated by projecting the summed allele frequencies to the US population using the Hardy-Weinberg equation and taking into consideration the X-linked mode of inheritance, assuming each individual can only carry 1 pathogenic variant.

It was found that pathogenic alleles are present in the gnomAD v4.1 sample for all variants in the non-Finnish European gnomAD ancestry group, for 2 variants in South Asian ancestry group, and for 1 variant in the African / African American and East Asian ancestry groups. For the remaining 5 ancestry groups, there are no pathogenic alleles recorded in the gnomAD v4.1 dataset across all 8 variants included for analysis in the study.

Results show the highest pathogenic allele carrier frequencies in the European (non-Finnish) ancestry group, followed by the South Asian, East Asian and African / African American ancestry groups. Using reported penetrance figures of 100% for males and 70% for females, it is estimated that the carrier and symptomatic populations of Fabry disease in the US in 2024, based on analysis of the 8 included variants, are **12**,**024** male carriers (or 1 in 14,022 males) who will all develop symptoms, and **24**,**845** female carriers (or 1 in 6,978 females), of whom 17,392 will develop symptoms. Of these carriers who will develop symptoms, around 98.6% (corresponding to 11,858 men and 17,153 women) will carry a variant primarily associated with late-onset or both forms of Fabry disease.

The prevalence figures presented in this study are significantly higher than those based on reported clinical cases and are in line with those presented more recently based on newborn screening studies and with the prevalence reported in the UK Biobank analysis. The US National Institute of Health reports Fabry disease prevalence at around 1 in 50,000 males (which would correspond to 1 in 25,000 females). Analysing just 8 of the potentially hundreds of causal variants within the *GLA* gene, this study suggests that Fabry disease may be over 3 times as prevalent as is currently believed. This work highlights the vast potential of large genetic databases to analyse rare diseases, which will continue to progress as these datasets add more data, which will improve their power and diversity.

**What Is Already Known On This Topic:**

- Fabry Disease is a rare X-linked lysosomal storage disorder with historical prevalence estimates ranging from 1 in 40,000 to 1 in 170,000 males, based on case ascertainment.
- More recent newborn screening studies that test alpha-galactosidase A activity or perform genetic testing within the *GLA* gene, in addition to a UK Biobank study examining the prevalence of selected causal Fabry disease variants, have consistently suggested that Fabry disease may be far more prevalent than the estimates based on case ascertainment.

**What This Study Adds:**

- To our knowledge, this is the first study providing population-level estimates of the number of causal Fabry disease carriers and of the symptomatic population in the US using publicly available data from gnomAD v4.1. Our estimates are consistent with those produced by newborn screening studies and the UK Biobank analysis, and suggest that late-onset Fabry disease may affect >1 in 10,000 people in the US in 2024 at some point during their lifetime.
- This study also demonstrates the potential of large genetic databases, such as gnomAD, for the study of rare genetic diseases, which are often misdiagnosed and may consequently be believed to be rarer than they are in reality.

**How This Study May Affect Research:**

- This study highlights two areas for improvement which would be significantly beneficial to the study of rare genetic diseases.
  ∘ While this study demonstrates the utility of genetic databases to study certain rare genetic diseases, it is likely that the study of rarer conditions, in particular those manifesting during childhood and/or with a dominant mode of inheritance, would be more difficult using genetic databases, as individuals with such conditions are less likely to be included in population-level genetic biobanks (such as UK Biobank) due to a healthy volunteer bias. It is important that future genetic datasets are more representative in their recruitment to ensure that rare genetic diseases are not systematically excluded or underrepresented among participants. Studies such as All Of Us in the US, and Our Future Health and the Generation Study in the UK, will be extremely helpful in addressing this point.
  ∘ Estimates of the symptomatic Fabry disease population in the US in 2024 were calculated using the most up-to-date penetrance estimates in males and females. However these estimates were calculated using individuals already present in a Fabry registry and therefore may overestimate the penetrance, and especially among females, since asymptomatic carriers may be less likely to join a disease registry. Accurate calculation of the symptomatic population with a given genetic disease relies upon accurate penetrance estimates, which are not always available. These estimates are best calculated from large population-level resources with linked genetic and electronic health record data.

## Introduction

Fabry disease is a rare genetic lysosomal storage disorder in which the body does not produce enough of the alpha-galactosidase A enzyme to break down sphingolipid molecules within the body, causing sphingolipid levels to build up to harmful levels in the blood vessels and organs, including the heart, kidneys and brain, eventually leading to potentially life-threatening symptoms including stroke, cardiovascular complications, and kidney failure [1, 2].

Fabry disease is caused by pathogenic variants within the *GLA* gene and, depending on the exact causal variant, may manifest as a severe early onset disease, referred to as classic Fabry disease, or a milder late-onset form. Epidemiological studies have produced varying estimates of Fabry disease prevalence (both forms combined); estimates based on reported clinical cases range from 1 in 40,000 to 1 in 170,000 individuals [3, 4], while more recent newborn screening studies have suggested a much higher prevalence of 1 in 1,250 to 1 in 21,973 individuals [5-8].

Fabry disease penetrance (the proportion of causal variant carriers who become symptomatic for the disease) has been estimated as 100% in males and 70% in females [9, 10] and, while the penetrance figure of 70% in females was an estimate published 20 years ago, both figures are still being cited as recently as 2022 [11]. More recently, a study analysing genetic data for 8 selected causal Fabry disease variants in the UK Biobank [12] found that 1 in 5,732 participants in that cohort carry a variant associated with the late-onset form of the disease. These variants were chosen by study investigators on the basis of being pathogenic / likely pathogenic, rare (present in <1 in 10,000 individuals), and either previously reported as causal for Fabry disease or protein-truncating.

In order to give an equivalent prevalence calculation for comparison, this study analysed the same 8 causal variants as those selected in the UK Biobank analysis, using publicly available data from gnomAD v4.1 [13] to provide estimates of the number of individuals in the US carrying a selected causal variant for Fabry disease, and of those individuals, the number that are expected to be symptomatic for Fabry disease at some point within their lifetime.

## Methods

### Allele Frequency Estimates

The 8 Fabry variants selected for inclusion in this analysis are presented in Table 1. In the previously published analysis using UK Biobank data [12], these variants were selected from a list of 81 pathogenic *GLA* coding variants as a result of being both rare and either previously reported as causing Fabry disease or having a protein truncating effect. Of these 8 variants, 7 are primarily associated with either the later onset form of the disease or both the later onset form and the classic form, and one variant is primarily associated with classic Fabry.

**Table 1.**
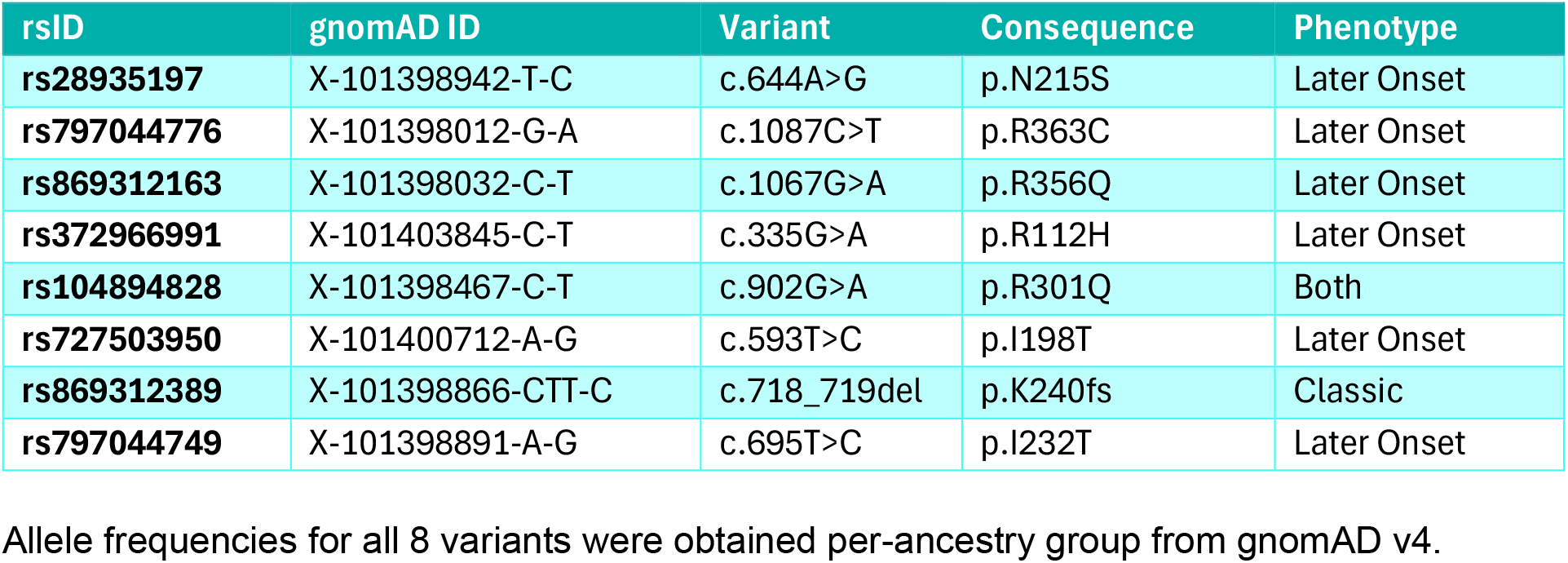
Fabry disease causal variants selected for inclusion in this study.

### Mapping US Population Figures to gnomAD Ancestry Groups

The size and demographic makeup of the US population in 2024 was obtained from the US Census Bureau [14]. Demographic data in the US Census is broken down by self-reported race featuring 5 main groupings, alone or in combination, while the global gnomAD v4.1 sample is broken down into genetically distinct ancestry groups. Some of these ancestry groups are very large and encompass many countries such as the African / African American and East Asian populations, while others are smaller and reflect finer-scale genetic differences within a wider ancestry group, such as the Ashkenazi Jewish, European (Finnish) and Amish populations, whose genetic architectures are from other groups of broadly European ancestry.

Population mapping and weighting between US Census groups and gnomAD ancestry groups is shown in Table 2 and reflects the proportion of gnomAD ancestry groups in each US Census group, for example 96.88% of the Non-Hispanic White Census group is of European (non-Finnish) ancestry. Weighting US Census Groups into multiple gnomAD ancestry groups was performed using previously published estimates of the proportions [15-18].

**Table 2.**
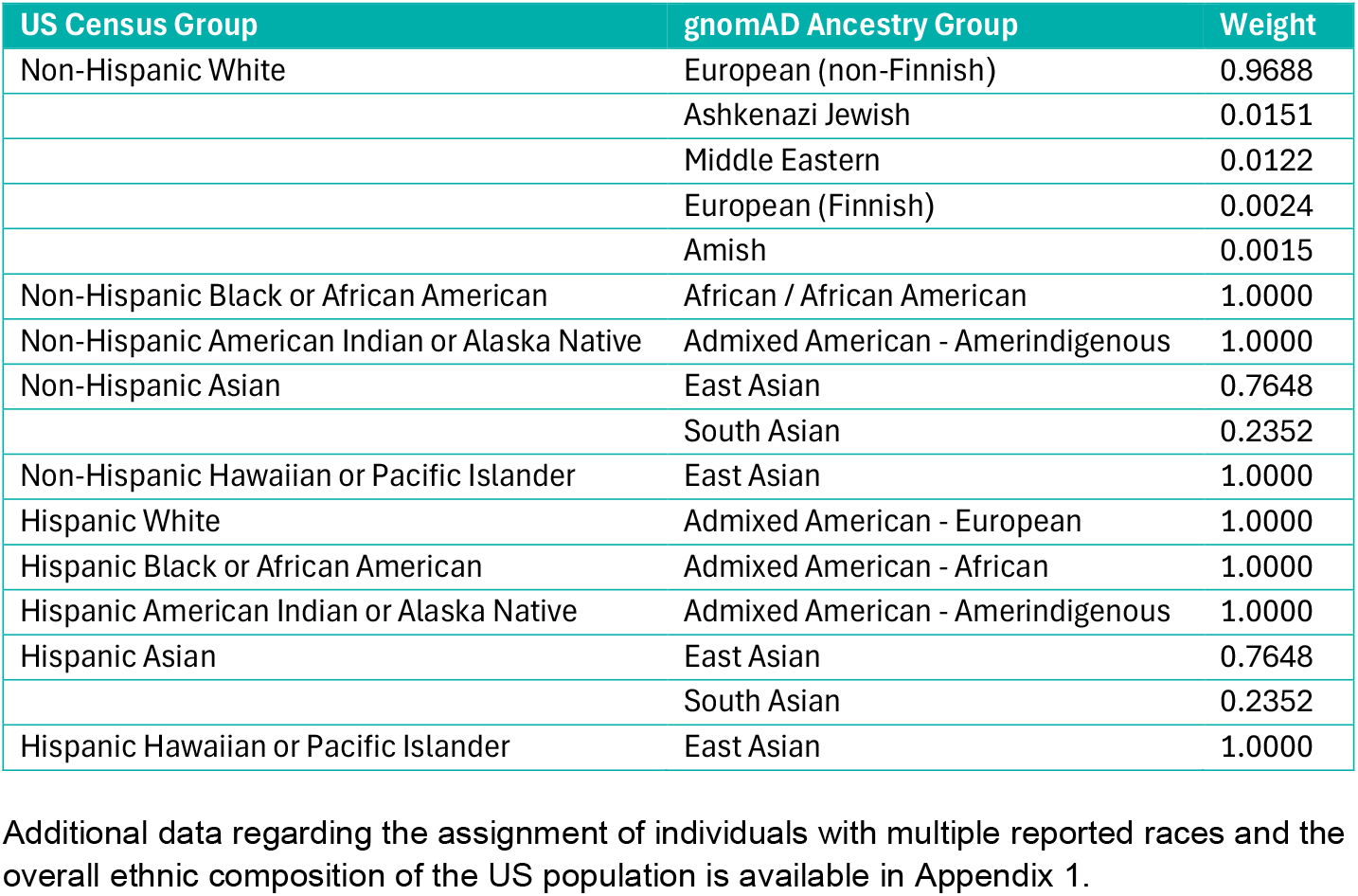
Mappings and weightings between US Census groups and gnomAD ancestry groups.

### Key Assumptions

This study used two sources of population data with different methods of defining subpopulations to disaggregate the data. The methods presented in this report are made with the following assumptions:

- American Indian or Alaska Native, Asian, and Hawaiian or Pacific Islander US Census groups are genetically equivalent in the Hispanic and non-Hispanic subsets of the US population.
- All age groups were considered equally in this study, so population numbers presented in the US population include the total age range in US Census data, from 0–110 years old.
- Allele frequencies from gnomAD can be converted to carrier frequencies in women using the Hardy-Weinberg equation, and in men by multiplying the population by the allele frequency. As all risk variants are X-linked, this assumes that:
  ∘ Men must inherit their copy of a risk variant from their mother.
  ∘ Women may inherit their copy of a risk variant from either of their parents, and both parents are equally likely to pass the variant on. This does not account for any potential reduction in reproduction among variant carriers of either sex as a result of having disease symptoms.
- Due to small sample sizes in gnomAD, variant allele frequencies are taken to be the same in males and females.
- Penetrance is assumed to be the same for all variants and in all ancestral groups.
- It is assumed that any individual will only carry a maximum of one pathogenic allele (i.e. there are no homozygote females or compound heterozygotes / hemizygotes).

## Results

### Allele Frequencies

Allele frequencies from gnomAD v4.1 for the 8 included genetic variants are displayed in Table 3. Pathogenic alleles are present in the gnomAD v4.1 sample for all variants in the non-Finnish European gnomAD ancestry group, for 2 variants in the South Asian ancestry, and for 1 variant in the African / African American and East Asian ancestry groups. For the remaining 5 ancestries (Ashkenazi Jewish, Middle Eastern, European (Finnish), Amish and Admixed American), the reported pathogenic allele frequencies are 0 for all included variants.

**Table 3.**
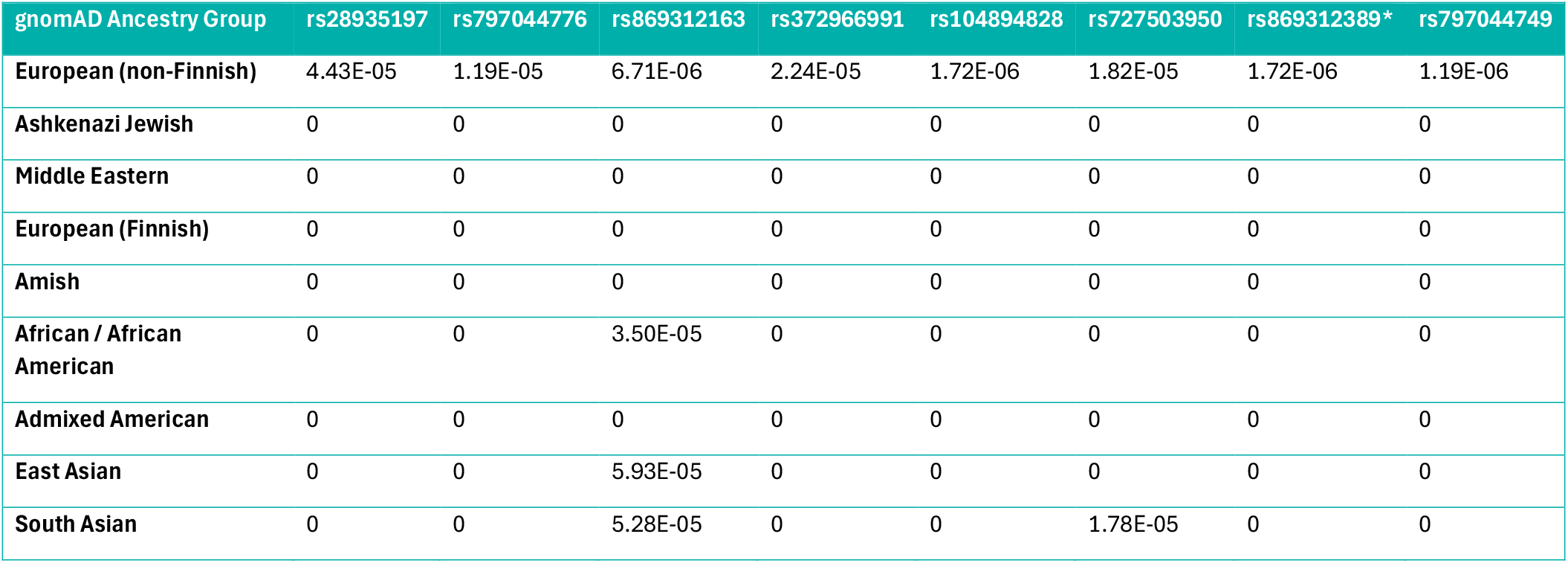
gnomAD v4.1 allele frequencies for all included variants. * variant primarily associated with classic Fabry.

### Carrier Population in the US: Women

The projected carrier population amongst women in the US is shown in Table 4. We calculate that an expected **24**,**845** or **1 in 6**,**978** women carry the pathogenic allele of one of the 8 genetic variants included in this analysis. Of these women, 24,504 or 1 in 7,075 carry a variant associated with either late-onset or both forms of Fabry disease.

**Table 4.** Calculated Fabry causal variant carrier populations and carrier rates amongst women in the US in 2024.

| US Census Group | gnomAD Ancestry Group | Weighted Population | Carriers | Carrier Rate |
| --- | --- | --- | --- | --- |
| **Non-Hispanic White** | **European (non-Finnish)** | 99,323,629 | 21,468 | 1 in 4,627 |
|  | **Ashkenazi Jewish** | 1,549,423 | 0 | - |
|  | **Middle Eastern** | 1,252,647 | 0 | - |
|  | **European (Finnish)** | 242,756 | 0 | - |
|  | **Amish** | 149,877 | 0 | - |
| **Non-Hispanic Black or African American** | **African / African American** | 23,444,050 | 1,640 | 1 in 14,295 |
| **Non-Hispanic American Indian or Alaska Native** | **Admixed American** | 1,792,091 | 0 | - |
| **Non-Hispanic Asian** | **East Asian** | 9,069,665 | 1,075 | 1 in 8,437 |
|  | **South Asian** | 2,789,207 | 394 | 1 in 7,079 |
| **Non-Hispanic Hawaiian or Pacific Islander** | **East Asian** | 480,735 | 57 | 1 in 8,434 |
| **Hispanic White** | **Admixed American** | 29,599,652 | 0 | - |
| **Hispanic Black or African American** | **African / African American** | 1,918,233 | 134 | 1 in 14,315 |
| **Hispanic American Indian or Alaska Native** | **Admixed American** | 1,124,555 | 0 | - |
| **Hispanic Asian** | **East Asian** | 361,902 | 43 | 1 in 8,416 |
|  | **South Asian** | 111,296 | 16 | 1 in 6,956 |
| **Hispanic Hawaiian or Pacific Islander** | **East Asian** | 155,438 | 18 | 1 in 8,635 |
| **All** | **All** | **173,365,156** | **24,845** | **1 in 6,978** |

### Carrier Population in the US: Men

The projected carrier population amongst men in the US is shown in Table 5. We calculate that an expected **12**,**024** or **1 in 14**,**022** men carry the pathogenic allele of one of the 8 genetic variants included in this analysis. Of these men, 11,858 or 1 in 14,218 carry a variant associated with either late-onset or both forms of Fabry disease.

**Table 5.** Calculated Fabry causal variant carrier populations and carrier rates amongst men in the US in 2024.

| US Census Group | gnomAD Ancestry Group | Weighted Population | Carriers | Carrier Rate |
| --- | --- | --- | --- | --- |
| **Non-Hispanic White** | **European (non-Finnish)** | 96,816,526 | 10,463 | 1 in 9,253 |
|  | Ashkenazi Jewish | 1,510,313 | 0 | - |
|  | Middle Eastern | 1,221,028 | 0 | - |
|  | European (Finnish) | 236,628 | 0 | - |
|  | Amish | 146,094 | 0 | - |
| **Non-Hispanic Black or African American** | **African / African American** | 21,666,883 | 758 | 1 in 28,584 |
| **Non-Hispanic American Indian or Alaska Native** | **Admixed American** | 1,717,738 | 0 | - |
| **Non-Hispanic Asian** | **East Asian** | 8,282,936 | 491 | 1 in 16,870 |
|  | South Asian | 2,547,263 | 180 | 1 in 14,151 |
| **Non-Hispanic Hawaiian or Pacific Islander** | **East Asian** | 484,688 | 29 | 1 in 16,713 |
| **Hispanic White** | **Admixed American** | 30,276,587 | 0 | - |
| **Hispanic Black or African American** | **African / African American** | 1,869,445 | 65 | 1 in 28,761 |
| **Hispanic American Indian or Alaska Native** | **Admixed American** | 1,194,295 | 0 | - |
| **Hispanic Asian** | **East Asian** | 356,675 | 21 | 1 in 16,985 |
|  | South Asian | 109,689 | 8 | 1 in 13,711 |
| **Hispanic Hawaiian or Pacific Islander** | **East Asian** | 162,506 | 10 | 1 in 16,251 |
| **All** | **All** | **168,599,294** | **12,024** | **1 in 14,022** |

### Carriers and Carrier Rate in gnomAD v4.1 Ancestry Groups

Figure 1 shows the carrier rates among men and women in the 4 gnomAD ancestry groups where at least one risk allele was present in the gnomAD sample. Carrier rates are highest in the European (non-Finnish) group, followed by South Asian, East Asian and African / African American ancestry groups.

**Figure 1.**
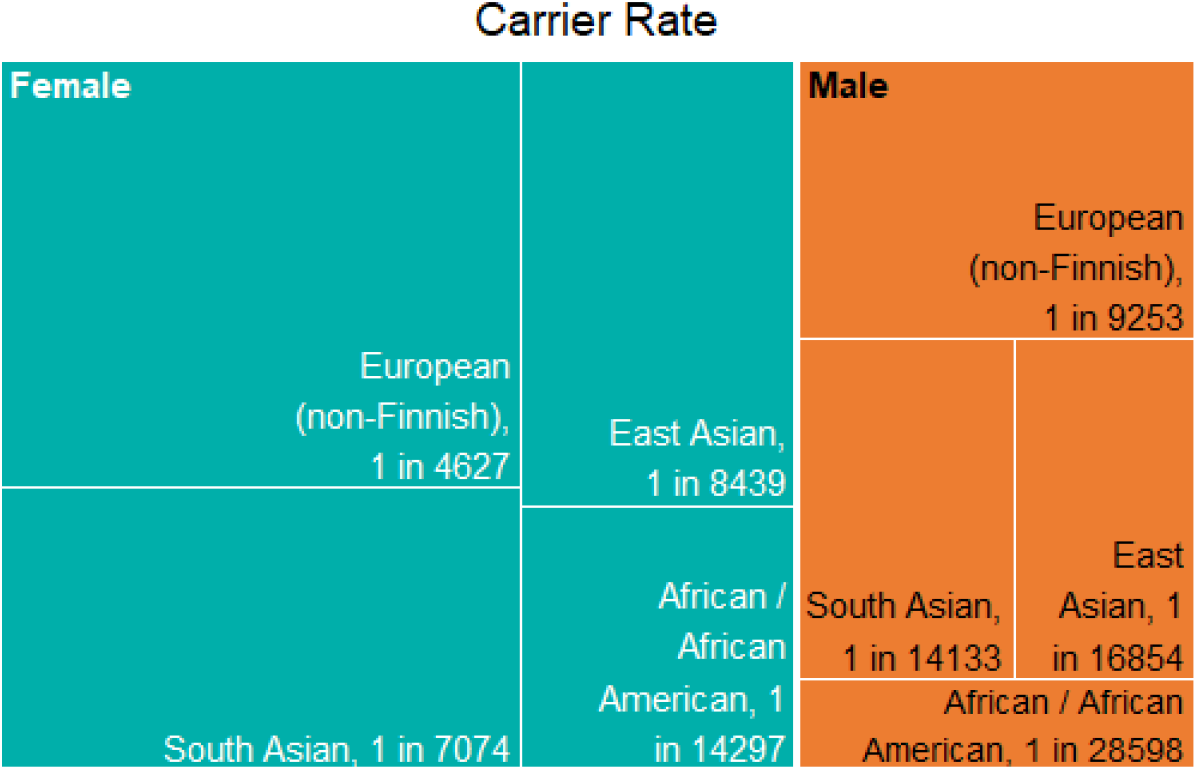
Treemap showing carrier rates in gnomAD genetic ancestry groups. Groups with a carrier rate of 0 (i.e. no risk alleles present) are not displayed.

### Symptomatic Population in the US: Lifetime Risk

Applying the published penetrance figures of 100% in males and 70% in females [9], we calculate that **12**,**024** males and **17**,**392** females in the 2024 US population will develop symptoms of Fabry disease at some point during their life. Of these, an expected 11,858 men and 17,153 women will carry a variant associated with late-onset or both forms of Fabry disease.

## Discussion

Using allele frequency data from gnomAD v4.1, an estimated 12,024 males and 24,845 females in the 2024 US population carry a risk allele for Fabry disease, of which around 98.6% (11,858 males and 24,504 women) carry one predominantly associated with late-onset or both forms of Fabry disease. These figures equate to approximately 1 in 14,000 males and 1 in 7,000 females in the US who carry one of the 8 variants included in this study.

The US National Institute of Health (NIH) reports Fabry prevalence of 1 in 50,000 males [19], which would correspond to 1 in 25,000 females. Similarly, the US Genetic and Rare Diseases Information Center (GARD) reports that fewer than 5,000 people in the US have Fabry disease [20]. The figures produced in this study, analysing just 8 of potentially more than 1,000 variants within the *GLA* gene [21], suggest that Fabry disease is over 3 times as prevalent as the NIH reports and, if the published penetrance estimates of 100% in males and 70% in females are accurate, there may be 6 times more cases in the US than GARD reports. The National Organization for Rare Disorders (NORD) reports that later-onset Fabry disease may be 5-10 times more prevalent than classic (juvenile onset) Fabry disease, which is consistent with the figures produced in this study [22].

A comparison of selected published Fabry disease prevalence estimates, in addition to this current study, is shown in Table 6. There is a clear distinction between prevalence estimates published on the basis of case studies, which report Fabry prevalence of <1 in 100,000 people, and those ascertained via newborn screening and genetic database analysis, which almost all report Fabry prevalence of >1 in 10,000 people, over an order of magnitude more prevalent than case study prevalence estimates. While there may be some uncertainty over the penetrance of the condition (the 70% female penetrance estimate was calculated using females registered in the Fabry registry and may therefore underrepresent females unknowingly living with a causal Fabry mutation [23]), multiple newborn screening studies have calculated prevalence estimates based on α-Gal A deficiency testing rather than genetic screening, with comparable findings. The majority of the Fabry causal variant carrier population in the UK Biobank had neither a Fabry disease diagnosis nor a medical history indicative of Fabry disease, suggesting that, among other possible explanations, the penetrance of these selected pathogenic variants is lower than current estimates suggest [12]. Penetrance estimates calculated using a sample more representative of the wider population may provide greater accuracy; however recruiting a sample size large enough to eliminate biases would prove difficult even in the context of a large biobank.

**Table 6.** Comparison of published Fabry disease prevalence estimates. VUS = variants of unknown significance.

| Authors | PMID | Published | Time Period | Location | Setting | Prevalence |
| --- | --- | --- | --- | --- | --- | --- |
| **Meikle *et al* [3]** | 9918480 | 1999 | 1st January 1980 to 31st December 1996 | Australia | Retrospective case studies | 1 in 117,000 |
| **Poorthuis *et al* [4]** | 10480370 | 1999 | 1970 to 1996 | The Netherlands | Enzymatically confirmed cases | 1 in 476,000 |
| **Spada *et al* [8]** | 16773563 | 2006 | 1st July 2003 to 30th June 2005 | Italy | Newborn screening (α-Gal A deficiency test and genetic test) | 1 in 3,100 males (α-Gal A deficiency and GLA mutation) |
| **Hwu *et al* [6]** | 19621417 | 2009 | July 2006 to June 2008 | Taiwan | Newborn screening (α-Gal A deficiency test) | ~1 in 1,250 males |
| **Burton *et al* [5]** | 28728811 | 2017 | 1st November 2014 to 31st August 2016 | Illinois, US | Newborn screening (α-Gal A deficiency test) | 1 in 8,454 |
| **Sawada *et al* [7]** | 31956509 | 2020 | August 2006 to December 2018 | Western Japan | Newborn screening (genetic test) | 1 in 11,854 (pathogenic variants), 1 in 7,683 (pathogenic plus VUS) |
| **Gilchrist *et al* [12]** | 35977816 | 2023 | 2006-2010 (UK Biobank recruitment) | UK | UK Biobank (genetic carriers of pathogenic Fabry variants) | 1 in 5,732 (late-onset variants), 1 in 200,643 (classic variants) |
| **Current study** | - | - | 2023-2024 (gnomAD v4.0 and v4.1) | US | gnomAD v4.1 projected to the 2024 US population (genetic carriers of pathogenic Fabry variants) | 1 in 14,022 males, 1 in 6,798 females |

This research used gnomAD v4.1 data to assess Fabry disease causal allele carrier rates in diverse genetic populations. However, the low sample sizes for some populations in gnomAD v4.1, combined with the rarity of the 8 variants included for analysis, results in an effect allele frequency of 0 for all 8 variants in these populations (Ashkenazi Jewish, Middle Eastern, European (Finnish), Amish and Admixed American). While it may be the case that Fabry disease is less common in those populations, it is unlikely to be completely absent and the lack of findings in this analysis is more likely to be a result of low power in the gnomAD v4.1 dataset. Future work will address this by using a more well-powered dataset including diverse populations, such as All Of Us in the US, which aims to recruit ∼1 million Americans from diverse backgrounds and ethnicities, including those who have previously been underrepresented in genomic research. This dataset was not complete at the time of analysis.

More broadly, this research highlights the utility of large genetic biobanks in the context of rare disease research. The sample included in the gnomAD v4.1 dataset is not selected to be representative of the wider population in terms of rare disease prevalence and may actually underrepresent many rare diseases, as individuals with severe pediatric disease and their first degree relatives are excluded from the dataset [24]. Additionally, over half of the gnomAD v4.1 sample is provided by UK Biobank, in which there is well-documented evidence of a healthy volunteer bias [25].

Despite these limitations, the Fabry disease prevalence reported in this study, calculated using gnomAD v4.1 allele frequencies and currently available penetrance estimates, far exceeds current prevalence estimates. However, it should be noted that the approach of using large genetic biobanks for rare disease research is likely best suited to the study of late-onset rare diseases, as was the case in this study, as patients with such conditions are less likely to be excluded from gnomAD. Findings from such analyses can be used to promote increased awareness of rare diseases among the medical community and provide pharmaceutical companies with more accurate estimates of the potential patient population size for therapeutic interventions.

## Data Availability

All data produced in the present work are contained in the manuscript

## Notes

### Competing Interest Statement

The authors have declared no competing interest.

### Funding Statement

This study did not receive any funding

### Author Declarations

The study used ONLY openly available human data that were originally located at https://gnomad.broadinstitute.org/

