## Supplemental Data 1 for "Estimating the prevalence of late-onset Fabry disease in the US in 2024"

### Supplementary 1

#### Assignment of Individuals with 2 or more Races

In the US Census individuals' ethnic and racial origin is recorded using two measures; racial origin (Non-Hispanic or Hispanic), and race code (White, Black or African American, American Indian and Alaska Native, Asian, Native Hawaiian and Other Pacific Islander, or 'Two or more races'). Each race code can be declared as either 'alone' (i.e. the individual is of one racial background), or 'in combination' where the individual has declared their racial background is 'two or more races'. Individuals declaring a single race code can be easily mapped to gnomAD ancestry groups as shown in Methods.

The subset of the population declaring multiple race codes in combination are mapped to gnomAD ancestry groups using the proportional representation of each race code within the subset of the US population checking 'two or more races' in their Census form, stratified by sex and racial origin (Hispanic or Non-Hispanic).

For example, 4,238,902 non-Hispanic men reported 'two or more races' on their Census return, with 8,841,945 race codes 'in combination' (shown in Table S1). Of those 8,841,945 race codes, 43.1% are 'White in Combination', 22.4% are 'Black or African American in Combination', 11.6% are 'American Indian and Alaska Native in Combination', 19.0% are 'Asian in Combination', and 3.9% are 'Native Hawaiian and Other Pacific Islander in Combination'. By applying these proportions to the 4,238,902 non-Hispanic men reporting 'two or more races', it is possible to estimate the racial makeup of this subset of the population, which can then be mapped to gnomAD ancestry groups in the same manner as those reporting a single racial origin.

Estimated populations for all 5 race groups in the Non-Hispanic and Hispanic origin groups are shown in Table S1 for men and Table S2 for women.

© HealthLumen Limited 2024 | Private and Confidential 

*Table S1. Assignment of racial origin to men with two or more reported races.*

| Origin | Race Code | Census Figures | Proportion | Estimated Population |
|---|---|---|---|---|
| **Not Hispanic** | **Total 'Two or more races'** | 4,238,902 | - | - |
| **Not Hispanic** | White in Combination | 3,812,744 | 0.431 | 1,827,855 |
| **Not Hispanic** | Black or African American in Combination | 1,983,247 | 0.224 | 950,782 |
| **Not Hispanic** | American Indian and Alaska Native in Combination | 1,021,318 | 0.116 | 489,627 |
| **Not Hispanic** | Asian in Combination | 1,677,995 | 0.190 | 804,442 |
| **Not Hispanic** | Native Hawaiian and Other Pacific Islander in Combination | 346,671 | 0.039 | 166,196 |
| **Hispanic** | **Total 'Two or more races'** | 1,106,248 | - | - |
| **Hispanic** | White in Combination | 994,442 | 0.419 | 463,523 |
| **Hispanic** | Black or African American in Combination | 530,968 | 0.224 | 247,492 |
| **Hispanic** | American Indian and Alaska Native in Combination | 442,330 | 0.186 | 206,176 |
| **Hispanic** | Asian in Combination | 304,046 | 0.128 | 141,720 |
| **Hispanic** | Native Hawaiian and Other Pacific Islander in Combination | 101,556 | 0.043 | 47,337 |

© HealthLumen Limited 2024 | Private and Confidential

*Table S2. Assignment of racial origin to women with two or more reported races.*

| Origin | Race Code | Census Figures | Proportion | Estimated Population |
|---|---|---|---|---|
| **Not Hispanic** | **Total 'Two or more races'** | 4,302,017 | - | - |
| **Not Hispanic** | White in Combination | 3,845,333 | 0.427 | 1,838,824 |
| **Not Hispanic** | Black or African American in Combination | 2,044,078 | 0.227 | 977,471 |
| **Not Hispanic** | American Indian and Alaska Native in Combination | 1,086,369 | 0.121 | 519,498 |
| **Not Hispanic** | Asian in Combination | 1,668,517 | 0.185 | 797,879 |
| **Not Hispanic** | Native Hawaiian and Other Pacific Islander in Combination | 352,042 | 0.039 | 168,345 |
| **Hispanic** | **Total 'Two or more races'** | 1,098,726 | - | - |
| **Hispanic** | White in Combination | 982,816 | 0.416 | 457,408 |
| **Hispanic** | Black or African American in Combination | 531,339 | 0.225 | 247,288 |
| **Hispanic** | American Indian and Alaska Native in Combination | 440,563 | 0.187 | 205,040 |
| **Hispanic** | Asian in Combination | 305,858 | 0.130 | 142,348 |
| **Hispanic** | Native Hawaiian and Other Pacific Islander in Combination | 100,217 | 0.042 | 46,642 |

© HealthLumen Limited 2024 | Private and Confidential

#### US Population Estimates

US population estimates by sex after assigning census groups to individuals of multiple reported racial origins are presented in Table S3.

*Table S3. US population estimates by sex after assigning census groups to individuals of multiple reported racial origins.*

| | Men | | | Women | | |
|---|---|---|---|---|---|---|
| **US Census Group** | **Single Race** | **Multi Race** | **Total** | **Single Race** | **Multi Race** | **Total** |
| **Non-Hispanic White** | 98,102,220 | 1,827,855 | 99,930,075 | 100,678,980 | 1,838,824 | 102,517,804 |
| **Non-Hispanic Black or African American** | 20,716,101 | 950,782 | 21,666,883 | 22,466,579 | 977,471 | 23,444,050 |
| **Non-Hispanic American Indian or Alaska Native** | 1,228,111 | 489,627 | 1,717,738 | 1,272,593 | 519,498 | 1,792,091 |
| **Non-Hispanic Asian** | 10,025,757 | 804,442 | 10,830,199 | 11,060,993 | 797,879 | 11,858,872 |
| **Non-Hispanic Hawaiian or Pacific Islander** | 318,492 | 166,196 | 484,688 | 312,390 | 168,345 | 480,735 |
| **Hispanic White** | 29,813,064 | 463,523 | 30,276,587 | 29,142,244 | 457,408 | 29,599,652 |
| **Hispanic Black or African American** | 1,621,953 | 247,492 | 1,869,445 | 1,670,945 | 247,288 | 1,918,233 |
| **Hispanic American Indian or Alaska Native** | 988,119 | 206,176 | 1,194,295 | 919,515 | 205,040 | 1,124,555 |
| **Hispanic Asian** | 324,644 | 141,720 | 466,364 | 330,850 | 142,348 | 473,198 |
| **Hispanic Hawaiian or Pacific Islander** | 115,169 | 47,337 | 162,506 | 108,796 | 46,642 | 155,438 |
| **All** | 163,253,630 | 5,345,150 | **168,598,780** | 167,963,885 | 5,400,743 | **173,364,628** |

© HealthLumen Limited 2024 | Private and Confidential 
